# Frequent Cocaine Use is Associated with Larger HIV Latent Reservoir Size

**DOI:** 10.1101/2022.03.31.22272986

**Authors:** Bradley E. Aouizerat, Josephine N. Garcia, Carlos V. Domingues, Ke Xu, Bryan C. Quach, Grier P. Page, Deborah Konkle-Parker, Hector H. Bolivar, Cecile D. Lahiri, Elizabeth T. Golub, Mardge H. Cohen, Seble G. Kassaye, Jack DeHovitz, Mark H. Kuniholm, Nancie M. Archin, Phyllis C. Tien, Dana B. Hancock, Eric Otto Johnson

## Abstract

**Background:** With the success of combination antiretroviral therapy, HIV is now treated as a chronic disease, including among drug users. Cocaine—one of the most frequently abused illicit drugs among persons living with HIV (PLWH)— slows the decline of viral production after ART, and is associated with higher HIV viral load, more rapid HIV progression, and increased mortality. We examined the impact of cocaine use on the CD4^+^ T-cell HIV Latent Reservoir (HLR) in virally suppressed PLWH.

**Methods:** CD4^+^ T-cell genomic DNA was isolated from peripheral blood mononuclear cells collected from 434 women of diverse ancestry (i.e., 75% Black, 14% Hispanic, 12% White) who self-reported cocaine use (i.e., 160 cocaine users, 59 prior users, 215 non-users). Participants had to have an undetectable HIV RNA viral load measured by commercial assay for at least 6 months. The Intact Proviral HIV DNA Assay (IPDA) provided estimates of intact provirus per 10^6^ CD4^+^ T-cells.

**Results:** The HLR size differed by cocaine use (i.e., median [interquartile range]: 72 [14, 193] for never users, for prior users 165 [63, 387], 184 [28, 502] for current users), which was statistically significantly larger in both prior (p=0.023) and current (p=0.001) cocaine users compared with never users.

**Conclusion:** Our study is the first to provide evidence that cocaine use may contribute to a larger replication competent HLR in CD4* T-cells among virologically suppressed women living with HIV. Our findings are important, because women are under-represented in HIV reservoir studies and in studies of the impact of cocaine use on outcomes among PLWH.

## Introduction

Combined antiretroviral therapy (ART) has successfully improved the life expectancy of people living with HIV (PLWH) and turned HIV into a chronic condition, managed by focusing on maintaining ART adherence, slowing progression, and preventing non-AIDS-defining illnesses.(*3*) The incidence of new HIV cases in the United States dropped to a relatively low 36,740 cases in 2019 overall and <3,000 cases among drug users in the United States.(*4*) Although isolated outbreaks of HIV highlight the need to maintain prevention efforts,(*5*) much of the HIV burden in the United States is among the more than 1 million PLWH,(*3, 4*) including among drug users(*4*).

Significant attention has shifted to finding a cure for HIV(*3, 6*), which is dependent on understanding and eliminating the HIV latent reservoir (HLR); i.e., the replication competent, but silenced, HIV provirus that is integrated into host cells’ DNA and is reactivated (i.e., begins to replicate) with cessation of ART.(*7*) Although consistent adherence to ART can reduce viral production to undetectable levels, no cure exists.(*8*) Discontinuing ART eventually results in activation of latent virus and resumption of viral production from functional latent virus.(*7-9*) A number of cell types are thought to harbor an HLR; the HLR residing in resting memory CD4+ T-cells(*8*) remains the best characterized and largest latent reservoir of silenced integrated HIV-1 proviruses.(*10, 11*) The HLR in resting CD4^+^ T-cells is initially established during acute infection(*12*) and added to during periods of non-adherence to ART(*13*). In virally suppressed subjects the HLR decays with a half-life of 44 months(*14, 15*), effectively precluding eradication with current therapies.

Cocaine (in either powder or crack form) is one of the most frequently abused non-prescribed drugs among PLWH, with current use estimates ranging from 10 to 30 percent.(*16-20*) With increasing engagement of people who use non-prescription drugs in HIV treatment,(*21*) it is likely that prevalence of cocaine use disorders among PLWH will grow even higher despite efforts to decrease cocaine use. Cocaine use is associated with accelerated HIV progression, which persists even with ART,(*1, 2*) and higher HIV viral load.(*22*) As reviewed by Dash *et al*.,(*23*) a preponderance of studies show adverse effects of cocaine use on HIV pathogenesis, including inferior virologic and immunologic responses to ART even among cART adherent patients(*24-29*) and higher likelihood of progressing to AIDS(*26, 30*). Of particular importance, Addai *et al*.(*31*) showed that administration of cocaine increases HIV-1 provirus integration in a dose-dependent manner and as much as 2.5-fold specifically in CD4+ T-cells. Given the persistent risk of relapse from cocaine abstinence(*32*), the contribution of cocaine use on the HLR could further delay the decay of the HLR. Additionally, a longer period of viral replication prior to ART suppression and higher pretreatment HIV viral load is associated with a larger HLR.(*33-35*) Collectively, these data suggest a mechanism by which cocaine use may associate with a larger HLR. Given these effects of cocaine use and the association of HIV viral load with HLR quantity,(*33-35*) cocaine use plausibly affects the HLR.

To our knowledge, only one study has examined the potential impact of cocaine use on the HLR.(*36*) Kirk *et al*.(*36*) investigated the associations of CD4^+^ T-cell HLR, quantified using the intact proviral DNA assay (IPDA)(*37, 38*), with active cocaine and/or heroin use in a subset of individuals with a history of injection drug use from the AIDS Linked to the IntraVenous Experience (ALIVE) Study. Although Kirk *et al*. found a slight trend toward increased HLR with active cocaine use only (p=0.18), the modest sample size (23 active cocaine only users, 28 no active illicit drug users) coupled with the complex drug use history of ALIVE participants, suggest that evaluation in a larger, independent sample is warranted. Moreover, the association of nonstructured ART treatment interruptions (i.e., periods of intermittent detectable viremia) as a proxy for ART nonadherence with a larger HLR by Kirk *et al*. (*36*), underscores the importance of accounting for the contribution of ART adherence history and suppression of HIV replication in the evaluation of the relationship of cocaine use on the HLR has not been done. In this study, we quantified the CD4^+^ T-cell HLR using the IPDA among ART adherent cocaine using (n=160) and ART adherent cocaine non-using (n=274) racially and ethnically diverse women living with HIV (WLWH) (i.e., 74.7% Black, 13.6% Hispanic, 11.8% White) in the Women’s Interagency HIV Study (WIHS) and tested whether alterations in the intact proviral HIV DNA reservoir were associated with cocaine use.

## Results

### Demographic, clinical, and substance use characteristics of study participants

In total, 544 HLR measures from 434 participants were evaluated. **Table 1** summarizes the characteristics of the cocaine use and non-use groups and the associations observed in bivariate analyses. Participants were all female, predominantly African American (75%), and 47 years old, on average (standard deviation [SD], 8.1 years). The mean of the most recent CD4^+^ T-cell count of the study participants was 706 cells/μL, and the mean nadir CD4^+^ T-cell count was 281 cells/μL. The median duration of virologic suppression was 1.5 years (interquartile range [IQR]: 0.5, 3.0). Compared to the non-cocaine using group, the cocaine use group reported more current cannabis use, more frequent high-risk drinking, and lower CD4^+^ T-cell count. Despite having undetectable viral load, a greater proportion of participants in the cocaine use group self-reported adhering to their ART regimen <95% of the time. There were no significant differences in mean age, self-reported race or ethnicity, tobacco use, or nadir CD4^+^ T-cell count by cocaine use.

**Table 1.**
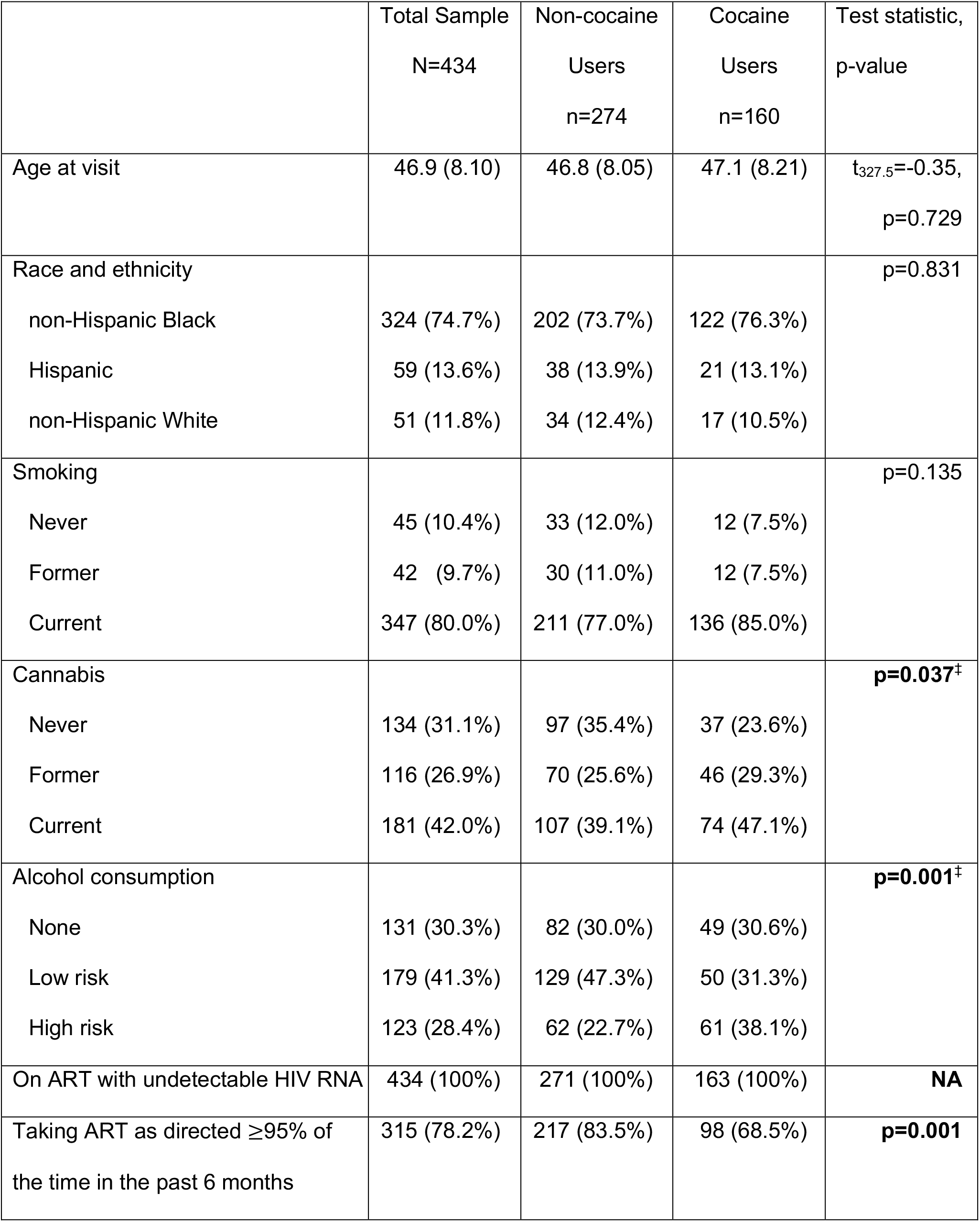
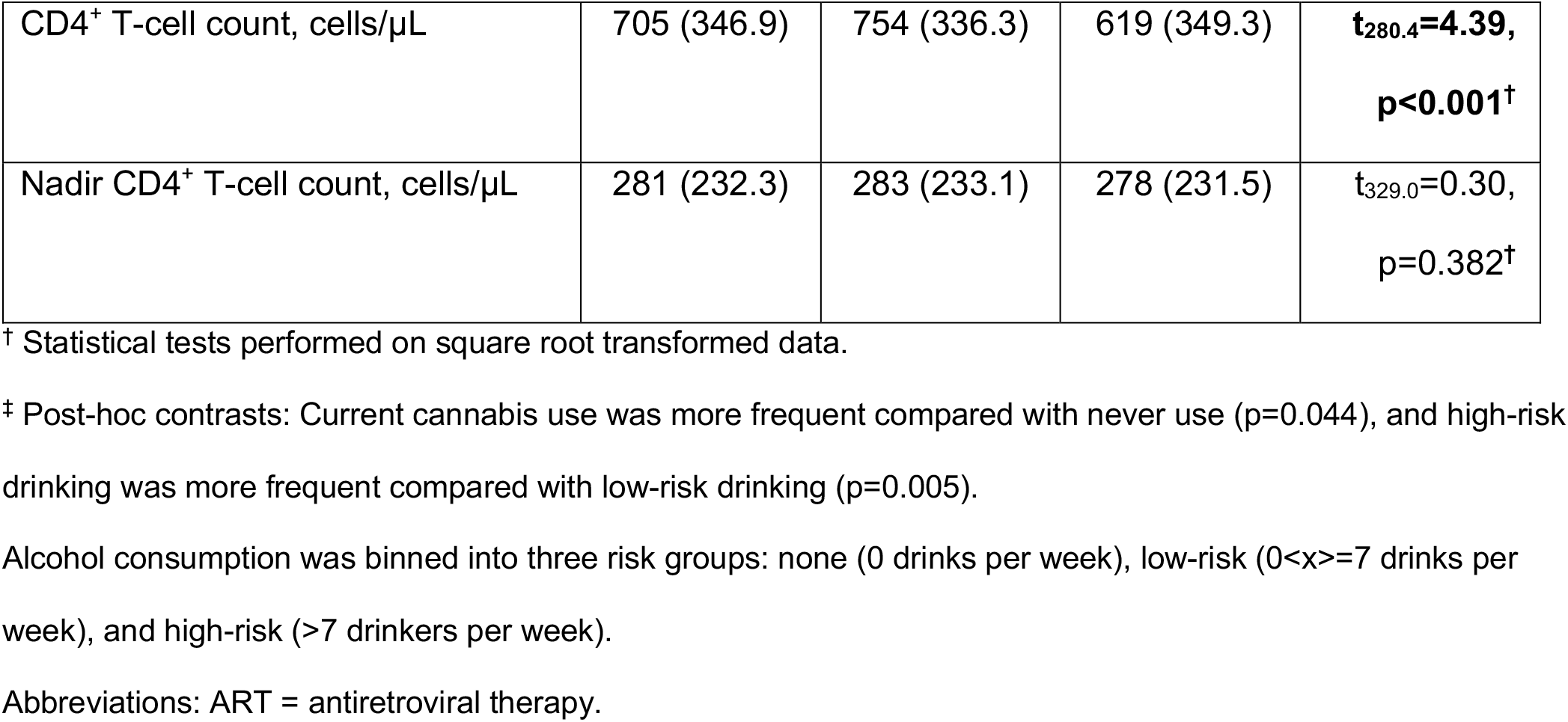
Demographic and Clinical Characteristics by Cocaine Use Group

### The impact of cocaine use on the HIV latent reservoir

Overall, the study sample displayed the expected distribution of intact and non-functioning proviral HIV DNA (**Figure 1**).(*39-41*) The intact proviral HLR was significantly larger in cocaine users (median [IQR]: 184 [28, 502]) as compared to non-users (median [IQR]: 87 [23, 262], p=0.0064). Violin plots of the distribution of intact proviral HIV DNA in active cocaine users and non-users is displayed in **Figure 2**. Among the 274 participants who reported not using cocaine in the prior 6 months, 59 participants reported use of cocaine in the past (median [IQR]: 4 years [1.5, 11.5]). While current cocaine use had the strongest effect on HLR size (median [IQR]: 72 [14, 193] for never users and 184 [28, 502] current users, n=375, p=0.001), participants who reported prior but not recent use of cocaine had a larger HLR but not statistically significant than women who never used cocaine (median [IQR]: 72 [14, 193] for never users and 165 [63, 387] for prior users, p=0.259)(**Figure 3**).

**Figure 1.**
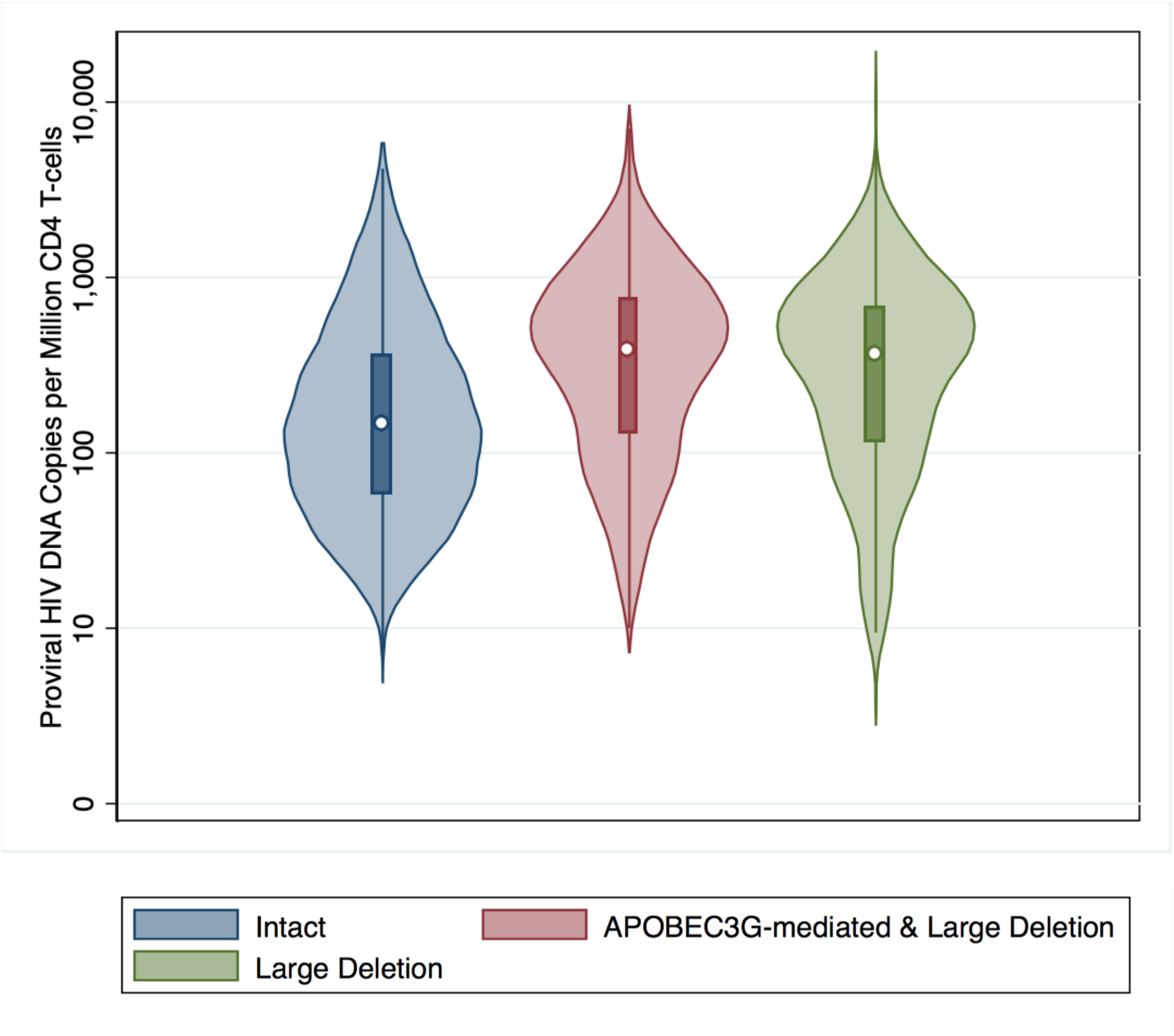
Box Plot of Intact and Mutated CD4^+^ T-cell HIV Proviral DNA Species. The IPDA assay measures intact (blue), APOBEC3G-mediated (red), and large deletion (red, green) proviral HIV genomes. As expected(*39-41*), the number of intact provirus was markedly lower than either class of non-functioning HIV proviral genomes, with the number of provirus harboring either large deletions or APOBEC3G-mediated mutations (also known as 3’ defective, red box plot) being more common than those harboring large deletions alone (also known as 5’ defective).

**Figure 2.**
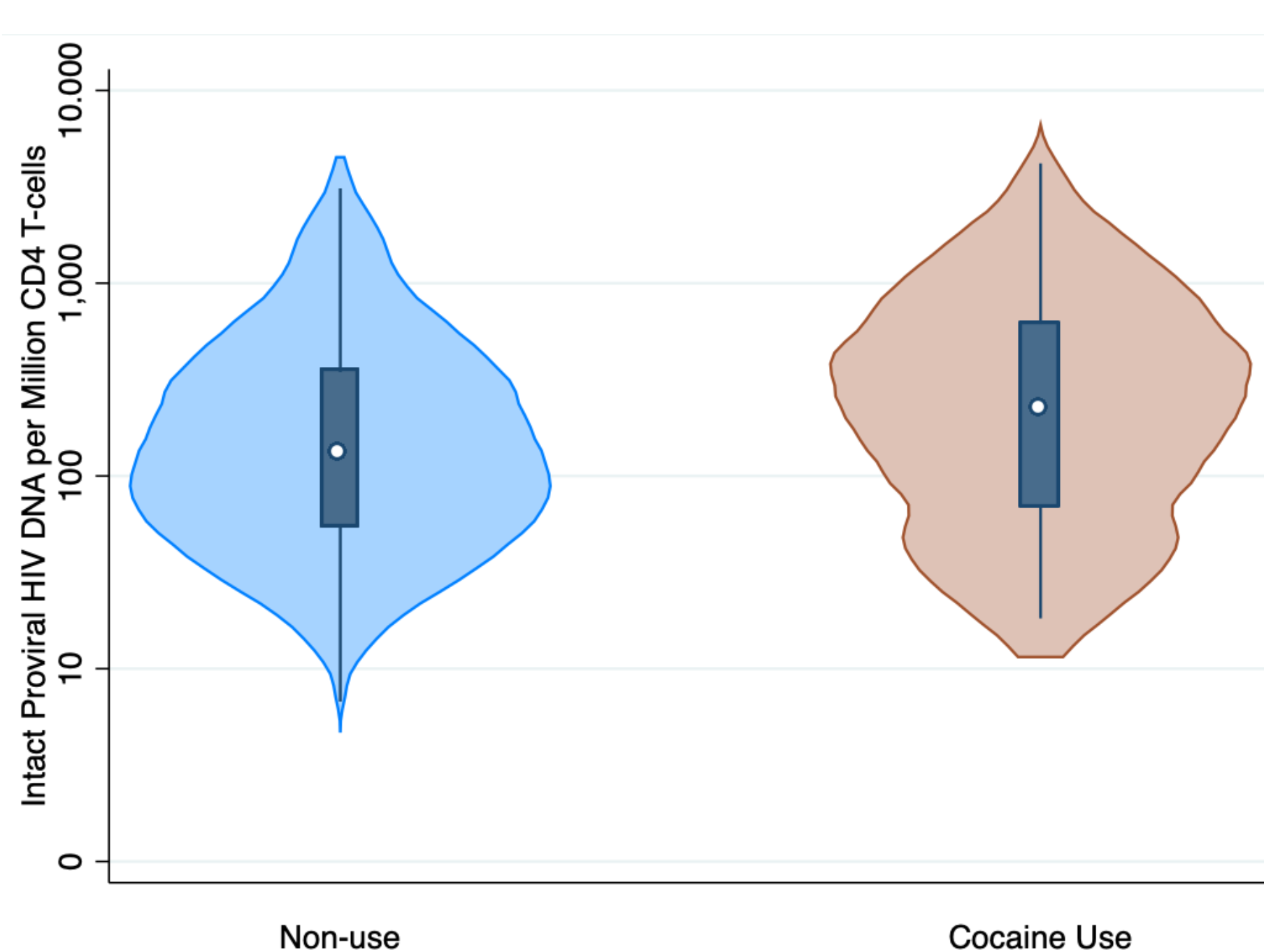
Violin Plot of Intact CD4^+^ T-cell proviral HIV DNA by Cocaine Use Group The number of intact proviral DNA per million CD4 T-cells was statistically significantly elevated in the Cocaine Use group (orange, n=160) as compared to the Cocaine Non-use group (blue, n=274). Generalized estimating equation Poisson regression model of the difference in intact proviral HIV DNA copies per million CD4^+^ T-cells in cocaine users as compared to non-users (p<0.001).

**Figure 3.**
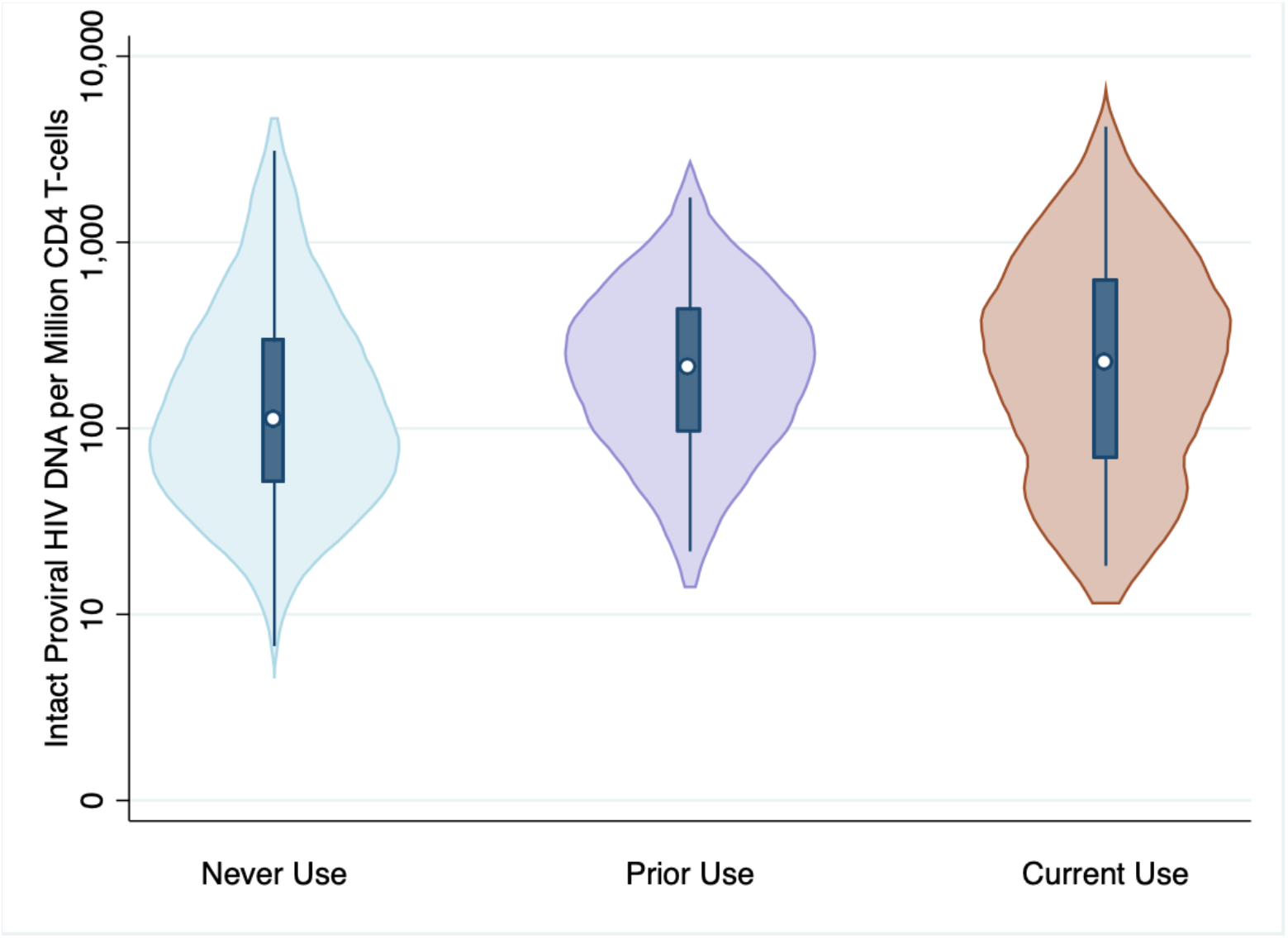
Violin Plot of Intact CD4^+^ T-cell Proviral HIV DNA by Never Use, Prior Cocaine Use, and Current Cocaine Use Groups. The median and interquartile range for the number of intact proviral HIV genomes per million CD4^+^ T-cells for each group was 72 (median): (14, 193) median (interquartile range [IQR]) for never users, 165 (63, 387) for prior users, and 184 (28, 502) for current cocaine users. Generalized estimating equation Poisson regression model of the difference in intact proviral HIV DNA copies per million CD4^+^ T-cells was statistically significantly elevated the Current Cocaine Use group (orange, n=160, p<0.001), but not in the Prior Cocaine Use group (n=59, p=0.259) as compared to the Never Use group (blue, n=215).

Because the factors that influence the HLR (e.g., past cocaine use, virological suppression, ART adherence) can vary over time, it is possible that sub-optimal self-management of HIV or use of cocaine could influence the HLR size. To address this possibility, we evaluated the impact of participants’ histories (collected every 6 months since enrollment) of cocaine use, virologic suppression, immune status (CD4^+^ T-cell count, nadir CD4^+^ T-cell count), and ART adherence using a repeated measures design on the HLR. We then evaluated whether participants’ current characteristics in terms of immune status (i.e., current CD4^+^ T-cell count – a measure of immune status in PLWH), treatment (ART adherence), or other substance use (i.e., alcohol consumption, cannabis use) influenced the relationship between cocaine use and the HLR (**Table 2**) using a GEE Poisson regression analysis. Current CD4^+^ T-cell count and self-reported ART adherence each attenuated the relationship between cocaine use and the HLR modestly; neither remained associated with HLR after controlling for cocaine use group when both exposures were included in the model. No evidence of residual confounding of alcohol use, smoking, or cannabis use on the relationship between cocaine use group and the HLR was observed (**Table 2**).

**Table 2.** Association of Cocaine Use, CD4 T-cell Count, and Antiretroviral Adherence on CD4^+^ T-cell Intact Proviral DNA HLR among Virally Suppressed Women.

|  | Beta<br>Coefficient | 95% CI | Z | p-value |
| --- | --- | --- | --- | --- |
| Unadjusted |  |  |  |  |
| Cocaine use vs non-use (ref) |  |  |  |  |
| Prior cocaine use | 2.09 | -1.210, 5.395 | 1.24 | 0.214 |
| Current cocaine use | 5.31 | 2.050, 8.569 | 3.19 | 0.001 |
| Adjusted for adherence |  |  |  |  |
| Cocaine use vs non-use (ref) |  |  |  |  |
| Prior cocaine use | 1.99 | -1.289, 5.260 | 1.19 | 0.235 |
| Current cocaine use | 4.57 | 1.340, 7.805 | 2.77 | 0.006 |
| Taking ART as directed <95% of the time in<br>the past 6 months | 2.97 | -0.757, 6.692 | 1.56 | 0.118 |
| Adjusted for CD4<sup>+</sup> T-cell count |  |  |  |  |
| Cocaine use vs non-use (ref) |  |  |  |  |
| Prior cocaine use | 1.56 | -1.639, 4.749 | 0.95 | 0.340 |
| Current cocaine use | 4.67 | 1.340, 8.009 | 2.75 | 0.006 |
| CD4<sup>+</sup> T-cell count | -0.23 | -0.566, 0.108 | -1.33 | 0.183 |
| Adjusted for alcohol consumption and cannabis use |  |  |  |  |
| Cocaine use vs non-use (ref) |  |  |  |  |
| Prior cocaine use | 1.90 | -1.401, 5.202 | 1.13 | 0.592 |
| Current cocaine use | 3.35 | 2.100, 8.601 | 3.23 | 0.001 |
| Alcohol consumption | -0.40 | -2.243, 1.450 | -0.42 | 0.647 |
| Cannabis use | -0.53 | -2.383, 1.320 | -0.56 | 0.573 |
| Tobacco use | 0.72 | -1.447, 2.888 | 0.52 | 0.515 |

General estimating equation Poisson regression models were fit to examine the associations between intact proviral HIV DNA at the index visit, adjusting for cocaine use group (current versus non-users) and CD4<sup>+</sup> T-cell count, cART adherence, alcohol consumption, cannabis use, and tobacco use. A
Abbreviations: ART = combined HIV antiretroviral therapy, CI = confidence interval, HLR = HIV Latent Reservoir.

When available, we also included repeated measures of the intact proviral HIV DNA reservoir (n=434, s=544). With the additional statistical power provided by repeated measures we observed that both prior and current cocaine use was associated with increases in HLR (**Table 3**). Additionally, prior detectable HIV viral load was associated with HLR *independent* of cocaine use. As expected, the association of HIV viral load history attenuated to the null following inclusion of history of ART adherence because these covariates are highly correlated. Longitudinal CD4^+^ T-cell counts was not associated with HLR when adjusting for cocaine use and history of HIV viral load. Self-reported race and ethnicity were not associated with the size of the HLR.

**Table 3.** Longitudinal Contribution of Cocaine Use and History of HIV Virologic Suppression on CD4^+^ T-cell Intact Proviral DNA HLR.

|  | Coefficient | 95% CI | Z | p-value |
| --- | --- | --- | --- | --- |
| Cocaine use vs non-use (ref) |  |  |  |  |
| Prior cocaine use | 2.66 | 0.364, 4.961 | 2.27 | 0.023 |
| Current cocaine use | 3.84 | 1.501, 6.176 | 3.22 | 0.001 |
| Historical HIV RNA levels | 6.82 | 5.984, 7.654 | 16.01 | <0.001 |

General estimating equation Poisson regression models were fit to account for the repeated measures of intact proviral HIV DNA, adjusting for cocaine use and HIV viral load. A log<sub>10</sub> transform was applied to normalize the distribution of HIV viral load.
Abbreviations: CI = confidence interval, HLR = HIV Latent Reservoir,

## Discussion

Our study shows for the first time that cocaine use may contribute to a larger replication competent HLR among virologically suppressed persons with HIV due to the increase in the CD4^+^ T-cell HLR. We found that cocaine users exhibited a significantly larger HLR as compared to non-users, including both current users and prior users. Cocaine use has been associated with progression of HIV despite virologic suppression(*23*), but the mechanism for this relationship remains undefined.

Our study focused on women, who are under-represented in studies of the HIV reservoir and the impact of cocaine use on outcomes among PLWH. A prior study of CD4^+^ T-cell HLR using the IPDA measure did not observe large differences in the reservoir occur based on sex(*34*), but the sample sizes were modest and cannot rule out more subtle differences in the HLR nor the impact of hormonal effects across the life course of women on the HLR. Moreover, these differences were not examined in relation to cocaine use. Given that cocaine use is independently associated with seroconversion among women(*42*), is more common in WLWH than their uninfected counterparts(*43*), and use persists even among ART-adherent persons(*23*), future studies aimed at evaluating sex differences, the contribution of hormonal effects across the life course, and cocaine use on the HLR in WLWH are warranted.

Our findings contrast with those from a recently published paper on a subset of the ALIVE Study(*36*), which showed no significant association between active cocaine use and HLR (p=0.18, but with a slight trend toward increased HLR with active cocaine use only). That study and ours both focused on CD4^+^ T-cells and used IPDA to quantify the HLR among suppressed participants with HIV in longitudinal studies. However, there are several study differences, most notably sample size and injection drug use history, that may explain the difference in findings. Kirk *et al*. included 108 participants with a history of injection drug use (23–29 people per group: no active illicit drug use, active heroin only, active cocaine only, active heroin and cocaine) whereas our study included 434 participants (274 non-cocaine and 160 cocaine using). The definitions of cocaine use differed between studies. We defined cocaine use as having used at least one time per week for at least the past 6 months, while non-users were composed of two groups: never users and prior users. Never users reported no cocaine use during their participation in WIHS, a 4- to 24-year timeframe. Prior users (i.e., more than 6 months prior to the index time point for the current study) included those who reported any cocaine use even if less than once per week or more in the past. Given that the HLR has a half-life of approximately 44 months(*14, 15*), a longer duration of cessation of cocaine use should result in diminishment of the HLR, a hypothesized effect that may explain the significant difference in the size of the HLR among prior cocaine users as compared to never users, but which was smaller than current cocaine users. Evaluating the duration of cessation and the amount of cocaine used in relation to HLR size warrants investigation in future studies. We also frequency balanced, to the extent possible, the cocaine use and non-use groups on age, tobacco use, cannabis use, and alcohol consumption. After adjusting for ART adherence, CD4^+^ T-cell count, and history of HIV viral load, we found that cocaine use remained strongly and significantly associated with HLR. In contrast, Kirk *et al*. used a less restrictive definition of users based on cocaine use in any amount in the past 6 months, and their controls were all people who have a lifetime history of injection drug use, who would have used heroin or cocaine in the past. Importantly, both Kirk *et al*. and our study found that nonstructured treatment interruptions (Kirk. et al.), which may be considered analogous to ART non-adherence (the present study), significantly contributed to HLR even among those that were virally suppressed, continuing to support the need for interventions to maintain treatment adherence among drug users. As expected, self-reported history of sub-optimal ART adherence, a proxy for HIV virologic suppression, was no longer significantly associated with HLR when history of HIV viral load was accounted for in the model, and was independent of the effect of cocaine use in our study.

Our study had some limitations. While the similarity in the estimates of the HLR between our sample of women and the sample of men and women of Kirk *et al*.(*36*) suggest that differences, if they exist, may be modest, a formal evaluation for sex differences in the relationship between cocaine use and the HLR is warranted. While approximately 20% of participants had HLR estimates below the limit of quantitation of 1 copy per million CD4^+^ T-cells, the proportion of samples below the limit of quantitation did not differ by cocaine use group (p=0.40). However, we cannot rule out that a subset of these estimates resulted from mutations in the integrated HIV provirus that interfere with primer annealing as demonstrated recently.(*44*) While we observed evidence of an effect of past use of cocaine on the HLR, our sample was not sufficiently large to estimate the impact of time since last use of cocaine or degree of cocaine use (i.e., exposure) on the HLR. Finally, while the IPDA assay detects intact provirus, it is likely to over-estimate the *functional* HLR, the degree to which is not currently known. Episodes of viremia resulting from cocaine use could reseed the HLR, increasing its size, but this plausible explanation is not yet tested. Studies have shown that cocaine use enhances HIV replication,(*45-47*) T-cell susceptibility to apoptosis,(*48*) and HIV provirus integration.(*31*)

Our results showed that cocaine use is associated with an increase in the HLR in virologically suppressed WLWH. Given the current state of the HIV epidemic, it is critically important to address the progression of HIV in cocaine users over the long term. Understanding the biological impacts of cocaine use on HLR, by leveraging omics data to assess gene regulation in relevant cell types, will take a vital step toward an HIV cure, particularly in the context of this unique population.

## Methods

### Study participants and samples

Using a nested case-control study design, participants were selected from the WIHS, a multi-center longitudinal observational cohort study of women living with or at risk for HIV infection (now known as the MACS-WIHS Combined Cohort Study).(*49*) Participants are representative of WLWH in the United States and underwent follow-up visits every six months which included the collection of peripheral blood mononuclear cells (PBMCs) and clinical, demographic, and substance use data. WIHS cohort characteristics, including details of the recruitment methods and enrollment, were described previously.(*50-52*)

With currently available assays, accurate HLR analysis requires at least a 6 month period of stable viremic suppression.(*34*) Accordingly, inclusion criteria included being on ART and having an undetectable HIV RNA viral load (i.e., below the limit of detection for the assay used at the time of testing) for at least 6 months prior to the study visit where substance use (i.e., cocaine, cannabis, tobacco, and alcohol) data and PBMCs were available for analysis. When available, samples from multiple timepoints from an eligible participant were retained for analysis. Cocaine use (cases) was defined as reported use of cocaine at least weekly during the last 6 months; non-use (controls) was defined as no cocaine use reported during the last 6 months. The groups were balanced by self-reported race and ethnicity (i.e., non-Hispanic African American/Black, Hispanic, non-Hispanic White) and age (5-year bins). To minimize potential confounding effects from other substance use, we balanced, to the extent possible with the participants available, the cocaine using and non-using groups on alcohol (i.e., non-use, low risk, high risk), tobacco (i.e., never, former, current), and cannabis (i.e., never, former, current) use over the past 6 months. Participants who reported use of other non-prescribed substances (e.g., heroin, methamphetamine, methadone, hallucinogens, “club drugs”) or those who reported less frequent cocaine use (i.e., less than once per week) at the eligible visit were excluded. A total of 434 treated WLWH were included in this case-control study (cocaine using participants (n)=160, measures (s)=165; non-cocaine using n=274, s=379). The number of repeated measures includes 54 participants with 2 measures, 18 participants with 3 measures, 4 participants with 4 measures, and 1 participant with 5 measures. Among the non-cocaine using group, 215 participants reported never using cocaine and 59 reported using cocaine in prior WIHS visits, but not in the 6 months prior to the visit included in this study.

### Measures

#### Clinical and demographic variables

Race, ethnicity, and age were collected by self-report. Use of ART was defined according to the Department of Health and Human Services Antiretroviral treatment guidelines.(*53*) ART adherence was defined dichotomously by participant self-report of taking their ART as instructed by their provider either ≥95% or <95% of the time over the last 6 months.(*54*) Peripheral blood CD4^+^ T-cell counts and HIV viral load quantitation in plasma were determined via standard assays in laboratories that participated in the NIAID laboratory quality assurance program. Nadir CD4^+^ was the lowest CD4^+^ T-cell count measured either as part of a participant’s follow-up in WIHS or prior to enrollment, which was ascertained via medical record review.

#### Substance use measures

Use of cocaine by any route (e.g., injection, nasal) or form (e.g., crack, powder), tobacco (e.g., cigarette, e-cigarette), and cannabis (e.g., combustion, ingestion) in the past 6 months were collected by self-report using the following 8 categories: never, less than once a month, at least once a month but less than once a week, once a week, 2–3 times a week, 4–6 times a week, once a day, and more than once a day. Alcohol consumption was collected by self-report as the number of drinks (defined as one serving of beer, wine, liquor, mixed drink) consumed per week and categorized as none (zero drinks per week), low risk (more than zero but less than 7 drinks per week), and high risk (more than 7 drinks per week or greater).(*55*) Cannabis and tobacco use was categorized as never, former, or current.

#### CD4^+^ T-cell HIV latent reservoir measured using the intact proviral HIV DNA assay

The IPDA assay detects intact and non-functional integrated HIV DNA (i.e., large deletions, APOBEC3G-mediated hypermutations).(*39*) CD4^+^ T-cells were isolated from 6–12 million PBMCs by negative bead selection using the DynaBeads Untouched Human CD4^+^ kit (ThermoFisher Scientific, Waltham, MA, catalog no. 11346D). Prior to CD4^+^ T-cell isolation, frozen viable PBMC aliquots were first thawed in a water bath, washed of DMSO with a cell medium (i.e., 10% heat inactivated fetal bovine serum (FBS) in RPMI), then treated with DNase I (Stem Cell Technologies, Cambridge, MA, catalog no. 07900) to decrease cell clumping and increase CD4^+^ T-cell yield. Purity of CD4^+^ T-cell isolation was initially verified in two samples by fluorescence-activated cell sorting, which showed purity of bead-based CD4^+^ T-cell isolation of 97.5%–99.0% (data not shown). Genomic DNA was isolated from CD4^+^ T-cells using magnetic bead-based nucleic acid isolation (Omega Biotek, Norcross, GA, catalog no. M6955) and spin column base isolation hand kits (i.e., Qiaamp DNA mini kit (Qiagen cat no. 51304). DNA was quantified by fluorometric assay (ThermoFisher Scientific, Waltham, MA, Qubit apparatus, catalog no. Q33230), DNA purity was assessed by spectrophotometry (ThermoFisher Scientific, Waltham, MA, NanoDrop apparatus), and DNA quality was determined using a microfluidic chip (Agilent, 2100 Bioanalyzer, catalog no. 5067-1508). DNA samples then underwent an additional bead removal step to remove any residual beads from upstream reactions as they can interfere with reaction droplet formation in droplet digital polymerase chain reaction (ddPCR). DdPCR was employed to measure the intact proviral HIV DNA using the modified IPDA assay as described by Bruner *et al*.(*39*), which provided estimates of intact provirus per 10^6^ CD4^+^ T-cells. CD4^+^ T-cell genomic DNA was employed as the input for IPDA, which was performed using the QX200 droplet Digital PCR System (Bio-Rad, Hercules, CA).

### Statistical Analyses

Data files were built and analyzed using Stata/SE 14.2 (StataCorp LLC, College Station, TX). Data were systematically examined for out-of-range values and data inconsistencies. Means and standard deviations, medians and interquartile ranges, or counts and proportions were used to describe the sample in terms of demographic, clinical, and treatment characteristics and CD4^+^ T-cell intact proviral HIV DNA levels.

A primary aim of the current study was to determine the effect of cocaine use on the size of the CD4^+^ T-cell HLR. Initially, Student’s *t*-test or Fisher’s exact test were performed to assess differences in demographic, clinical, and treatment characteristics between cocaine users and non-users (the reference group). Nonparametric tests (i.e., Kruskil-Wallace [current cocaine users, prior cocaine users, never users] and Wlcoxon [current cocaine users, non-users] tests) were performed to assess differences in the HLR between cocaine users and non-users (the reference group). Characteristics that differed between the two groups in bivariate analyses (p<0.05) were evaluated as covariates in subsequent regression models. Generalized estimating equations (GEE) using a Poisson distribution were employed to evaluate the association of cocaine use with the CD4^+^ T-cell HLR to account for its non-normal distribution with a large number of zeros, adjusting for relevant covariates (e.g., while accounting for history of ART adherence, immune status [current and nadir CD4^+^ T-cell count], historical HIV viral load) and the repeated measures available in some participants. Participant was used as the clustering variable. Even though all participants had an undetectable HIV viral load in the prior 6 months, history of treatment adherence was included as a covariate to account for previous episodes of suboptimal adherence that may influence the size of the HLR. Tests with a *p-*value of <0.05 were considered statistically significant. Post-hoc contrasts accounted for multiple testing by applying a Bonferroni correction for the number of tests.

### Study approval

Informed consent was provided by all participants via protocols approved by institutional review committees at each affiliated institution.

## Data Availability

Data may be obtained through the MACS/WIHS Combined Cohort Study collaboration and Concept Sheet process.

https://statepi.jhsph.edu/mwccs/work-with-us/

## Author contributions

Conceptualization was contributed by BEA and EOJ. Methodology was contributed by BEA, JNG, CVD, and EOJ. Formal analysis was contributed by BEA. Resources were contributed by BEA and EOJ. Writing of the original draft was contributed by BEA, EOJ, DBH, BCQ, and KX. Review and editing were contributed by BEA, EOJ, DBH, BCQ, KX, GPP, DK-P, HHB, CDL, ETG, MHC, SGK, JD, MHK, NMA, and PCT. Funding acquisition was contributed by BEA and EOJ.

## Acknowledgments

The contents of this publication are solely the responsibility of the authors and do not represent the official views of the National Institutes of Health (NIH). Data collection and the contributions of Bradley E. Aouizerat, Josephine N. Garcia, Carlos V. Domingues, Ke Xu, Bryan C. Quach, Grier P. Page, Dana B. Hancock, and Eric Otto Johnson were supported by R66 DA047011, R66 DA047011-S1, and R33 DA047011. Clinical data and specimens used in this manuscript were collected by the Women’s Interagency HIV Study (WIHS), now the MACS/WIHS Combined Cohort Study (MWCCS). MWCCS (Principal Investigators): Atlanta CRS (Ighovwerha Ofotokun, Anandi Sheth, and Gina Wingood), U01-HL146241; Bronx CRS (Kathryn Anastos and Anjali Sharma), U01-HL146204; Brooklyn CRS (Deborah Gustafson and Tracey Wilson), U01-HL146202; Data Analysis and Coordination Center (Gypsyamber D’Souza, Stephen Gange and Elizabeth Golub), U01-HL146193; Chicago-Cook County CRS (Mardge Cohen and Audrey French), U01-HL146245; Northern California CRS (Bradley Aouizerat, Jennifer Price, and Phyllis Tien), U01-HL146242; Metropolitan Washington CRS (Seble Kassaye and Daniel Merenstein), U01-HL146205; Miami CRS (Maria Alcaide, Margaret Fischl, and Deborah Jones), U01-HL146203; UAB-MS CRS (Mirjam-Colette Kempf, Jodie Dionne-Odom, and Deborah Konkle-Parker), U01-HL146192; UNC CRS (Adaora Adimora and Michelle Floris-Moore), U01-HL146194. The MWCCS is funded primarily by the National Heart, Lung, and Blood Institute (NHLBI), with additional co-funding from the Eunice Kennedy Shriver National Institute Of Child Health & Human Development (NICHD), National Institute On Aging (NIA), National Institute Of Dental & Craniofacial Research (NIDCR), National Institute Of Allergy And Infectious Diseases (NIAID), National Institute Of Neurological Disorders And Stroke (NINDS), National Institute Of Mental Health (NIMH), National Institute On Drug Abuse (NIDA), National Institute Of Nursing Research (NINR), National Cancer Institute (NCI), National Institute on Alcohol Abuse and Alcoholism (NIAAA), National Institute on Deafness and Other Communication Disorders (NIDCD), National Institute of Diabetes and Digestive and Kidney Diseases (NIDDK), National Institute on Minority Health and Health Disparities (NIMHD), and in coordination and alignment with the research priorities of the National Institutes of Health, Office of AIDS Research (OAR). MWCCS data collection is also supported by UL1-TR000004 (UCSF CTSA), UL1-TR003098 (JHU ICTR), UL1-TR001881 (UCLA CTSI), P30-AI-050409 (Atlanta CFAR), P30-AI-073961 (Miami CFAR), P30-AI-050410 (UNC CFAR), P30-AI-027767 (UAB CFAR), and P30-MH-116867 (Miami CHARM).

The authors gratefully acknowledge the contributions of the study participants and dedication of the staff at the MWCCS sites.

